# The Course of Relapse Following Antipsychotic Discontinuation in Schizophrenia: A Test of the Antipsychotic Withdrawal Syndrome Hypothesis of Relapse

**DOI:** 10.64898/2026.01.19.26344378

**Authors:** Krisya Louie, Sameer Jauhar, Jose Rubio, Bodyl Brand, Toby Pillinger, Oliver D. Howes, Robert A. McCutcheon

## Abstract

**Background:** Antipsychotics are central to relapse prevention in schizophrenia, but longer-term use is associated with adverse effects that often prompt dose reduction or discontinuation. Although relapse risk increases after discontinuation, the nature of relapse remains unclear. Specifically, it is uncertain whether relapse reflects re-emergence of underlying illness or pharmacological withdrawal.

**Methods:** We analysed longitudinal symptom data (Positive and Negative Syndrome Scale; PANSS) from 417 individuals with schizophrenia who experienced relapse post-stabilization in five randomized, double-blind, placebo-controlled discontinuation trials of oral and long-acting injectable (LAI) paliperidone. Latent class mixed modelling was used to identify distinct trajectories of symptom change preceding relapse.

**Findings:** Two latent classes of relapse were identified: ‘rapid’ and ‘delayed’ onset. Rapid relapse was associated with more severe positive, negative, and cognitive symptoms at relapse. The proportion of individuals experiencing rapid relapse did not differ between those randomized to placebo (treatment discontinuation) versus treatment continuation in either LAI (p=0.119) or oral trials (p=0.949). No consistent increase in withdrawal-like symptoms (e.g. anxiety, agitation, depression) was found in discontinuation compared to continuation groups. Across formulations, individuals with rapid relapse had significantly higher baseline PANSS scores than those with delayed relapse (p<0.001).

**Interpretation:** Relapse following antipsychotic discontinuation follows at least two distinct trajectories that are not specific to treatment withdrawal. The comparable proportions of rapid and delayed relapse trajectories between discontinuation and continuation groups, together with the absence of a distinct symptom profile at relapse, does not support pharmacological withdrawal as a common mechanism of relapse. Instead, higher baseline symptom severity in those who experience rapid relapse may reflect pre-existing vulnerability and/or trial-related measurement artefacts related to baseline symptom rating and trial inclusion criteria. This emphasizes the clinical importance of risk stratification and individual monitoring, and challenge the assumption that relapse risk can be meaningfully reduced through dose tapering strategies alone.

## Introduction

Schizophrenia is a severe psychiatric disorder which often shows a relapsing-remitting illness course.^1^ Relapse is common and associated with marked functional disruption^2^, making relapse prevention a primary objective of long-term illness management.^3^ Antipsychotic medications are the foundation of maintenance treatment, which primarily exert their therapeutic effects through dopamine D_2_ receptor antagonism.^4^ Meta-analyses of randomized controlled trials (RCTs) have demonstrated the efficacy of antipsychotics in reducing relapse risk compared to placebo,^5,6^ supporting clinical guidelines that recommend maintenance therapy in individuals at risk of recurrence, particularly during the early phase of illness.^7,8^

Despite their efficacy, long-term antipsychotic treatment is associated with substantial side effects, which can impair quality of life and functional recovery, leading some people to request dose reduction or discontinuation.^9,10^ Side-effects and lack of insight contribute to non-adherence, which is one of the main factors associated with relapse.^11^ With increasing emphasis on shared decision-making and patient autonomy, there has been growing interest in whether antipsychotic medication can be safely reduced or discontinued in individuals who have achieved sustained symptom stability.^12,13^ Current guidance endorses individualized treatment management strategies that balance relapse risk against tolerability.^14,15^ However, at a group level, the risk of relapse after antipsychotic discontinuation is approximately twice those observed with continued maintenance treatment.^5^ There remains a paucity of methodologically robust evidence to guide clinicians on how best to discontinue antipsychotics, how relapse should be monitored during discontinuation, and whether particular symptom patterns are early warning signs of relapse.^16^

Competing hypotheses have been proposed to explain the nature of relapse following antipsychotic discontinuation (eTable 1). The conventional view holds that relapse reflects re-emergence of underlying illness once dopamine D_2_ receptor blockade drops below the effective threshold.^17^ This interpretation is supported by meta-analyses finding no significant difference in risk of relapse between abrupt and gradual antipsychotic discontinuation,^5,18^ suggesting that relapse is primarily driven by insufficient dopamine receptor blockade rather than the speed of dose reduction.^19^ An alternative hypothesis proposes that some cases of relapse represent withdrawal phenomena such as dopamine supersensitivity psychosis, particularly in rapid-onset cases accompanied by new antipsychotic withdrawal symptoms.^20^ Here, chronic D_2_ blockade induces compensatory upregulation of D_2_ receptors, increasing post-synaptic sensitivity to endogenous dopamine and triggering rebound symptoms upon drug discontinuation.^21^ While this hypothesis is supported by rat models and some clinical observations, its prevalence and generalizability remain uncertain.^22–24^

Some antipsychotic discontinuation trials report a disproportionately large number of people with early relapse, which has been interpreted by some as evidence for withdrawal effects.^16,24^ However, an alternative explanation for this observed rapid relapse patterns involves methodological artefacts in clinical trials.^19^ Recruitment pressures can create incentives for symptom ratings to be artificially lowered during the pre-randomization phase to meet eligibility thresholds.^25^ Once randomized, these pressures are relaxed and subsequent symptom ratings may more accurately reflect clinical status, creating the appearance of abrupt deterioration, leading to interpretation of an apparent abrupt relapse. These competing hypotheses raise the possibility that relapse may not be a single phenomenon, but might represent multiple trajectories, with different underlying neurobiology and symptom dynamics.

Resolving this debate is complicated by limitations of conventional analytical methods. Most RCTs employ time-to-relapse analyses, using Kaplan-Meier survival models to estimate the hazard of relapse.^18^ While appropriate for quantifying *when* relapse occurs, survival analyses treat relapse as a single binary outcome, which may obscure *how* symptoms evolve over time, leading up to relapse. In clinical practice, patients may follow different illness courses prior to relapse: some may have illnesses that deteriorate rapidly, others more slowly or intermittently. A key question is whether relapse after antipsychotic discontinuation differs in trajectory and symptomatology from relapse occurring during continued treatment (“breakthrough” relapse).

To address this limitation, we applied latent class mixed modelling (LCMM) to longitudinal symptom data from randomized antipsychotic discontinuation trials. LCMM is a data-driven method that identifies latent subgroups of individuals with similar trajectories of symptom change.^26,27^ This analytic approach enables the identification of distinct relapse trajectories, such as rapid-onset or delayed patterns.

Our main aim was to test whether the ratio of trajectory classes (e.g. rapid compared to delayed) was greater in the discontinuation arm compared to the continuation arm of randomized discontinuation trials, as this would support a pharmacological withdrawal form of relapse. Alternatively, hypotheses involving baseline score deflation and underlying illness-related processes, would predict similar distributions of relapse trajectories across treatment groups. In addition, we aimed to investigate symptom profiles at the time of relapse using item-level Positive and Negative Syndrome Scale (PANSS) scores^28^ to evaluate whether symptoms plausibly related to withdrawal (e.g., agitation, anxiety, depression) were more prominent in certain trajectory classes or treatment groups. Finally, we planned to evaluate baseline symptom profiles to examine whether rapid relapses are associated with greater severity of symptoms at time of randomization – supporting the baseline inflation hypothesis.

## Methods

### Identification of study cohort and data extraction

The Yale Open Data Access (YODA)^29^ was searched for randomized discontinuation trials of antipsychotics in individuals aged 18-65 with schizophrenia or schizoaffective disorder. Double-blind placebo-controlled relapse prevention RCTs of antipsychotic medications were included. In these trials, participants were first treated with antipsychotic drugs for more than three months and clinically stabilized before they were randomized to placebo (discontinuation) or continued active treatment. Five eligible trials were identified with available individual participant data examining both oral and long-acting injectable (LAI) formulations of paliperidone palmitate versus placebo.

The cohort of interest was defined as participants that experienced relapse regardless of treatment group at any time post-randomization during the double-blind phase according to the original study criteria (Supplementary Material 3). The individual participant data analyzed in this study included: treatment group, age, sex, baseline Clinical Global Impression (CGI) scores^30^ and individual item scores on the Positive and Negative Syndrome Scale (PANSS)^28^ at baseline and at all available measurement timepoints from the date of randomization up until date of relapse. The time to relapse was calculated as the number of days elapsed from the date of randomization.

### Statistical analysis

LCMM was employed to identify subgroups with distinct trajectories of psychotic (PANSS) symptoms among individuals who experienced illness relapse during ongoing antipsychotic treatment or after treatment discontinuation. LCMM is a statistical modelling technique that can analyze the change in longitudinal outcomes over time, and identify latent classes with distinct outcome trajectories.^31^ LCMMs are an extension of linear mixed models developed to handle population heterogeneity through uncovering unobserved (latent) classes of trajectories, based on longitudinal repeated measures.^27^ Unlike conventional mixed-effects models that assume a single population symptom trajectory, LCMM can identify distinct trajectory classes by simultaneously modelling the pattern of within-person change, and between-person variability. By taking a data-driven approach to derive latent classes of symptom patterns, LCMM avoids incorporating a priori assumptions of group structure. This analytical method is well-suited to studying complex fluctuating latent processes, such as the experience of psychotic symptoms over time, as it can detect subtle individual differences in symptom progression and capture patterns of clinical variability leading up to illness relapse.^26^

The study cohort was split into two groups: an oral formulation cohort and an LAI cohort, for model fitting and subsequent analyses. A secondary analysis further splitting the LAI cohort into 1-monthly and 3-monthly formulations was also conducted. This approach allowed more optimal model fit and comparison between different formulations of paliperidone, with varying half-lives and pharmacokinetic profiles. A series of latent class models were fitted on the total PANSS scores from all available measurement timepoints, starting from initial randomization visit up to the date of relapse, irrespective of treatment status. An iterative process to model specification within the maximum likelihood framework was implemented (Supplementary Material 1). Goodness-of-fit of the selected model was confirmed by plotting weighted subject-specific predicted trajectories against observed values for each latent class.

After selecting the final optimal latent class model, each individual was assigned to the latent trajectory class with the highest posterior probability of membership. Chi-square tests of independence were performed to evaluate whether the proportion of individuals in specific relapse trajectory classes differed between treatment groups. Symptom profiles in terms of individual PANSS item scores at time of relapse and at baseline were compared between latent trajectory classes and between treatment groups, including symptoms previously proposed to reflect withdrawal rather than true psychotic withdrawal, such as anxiety, depression and agitation (see eTable 1). Differences between latent classes and between treatment groups in covariates of interest (original study trial, sex, age, baseline PANSS and CGI scores) were assessed with the appropriate statistical test, with correction for multiple testing (Supplementary Material 1). LCMM was conducted with the R package *lcmm*.^31^ All statistical analyses were conducted in R version 4.3.0^32^ on the secure YODA platform.^33^

## Results

The final study cohort consisted of 417 participants who relapsed during the double-blind post-randomization phase of the five antipsychotic discontinuation trials, with 271 in the three long-acting injectable (LAI) trials and 146 in the two oral formulation trials (eTable 2). More individuals in the discontinuation group experienced relapse (LAI:72.7%; oral:74.0%) compared to continued active treatment (LAI:27.3%; oral:26.0%).

For both LAI and oral formulation groups, the two-class linear transformation latent class mixed model with random effects had the best fit, according to the predefined statistical criteria (eTable 3). There was general agreement between the predicted values and actual observations in the weighted mean subject-specific prediction plots by latent class (Figure 1).

**Figure 1.**
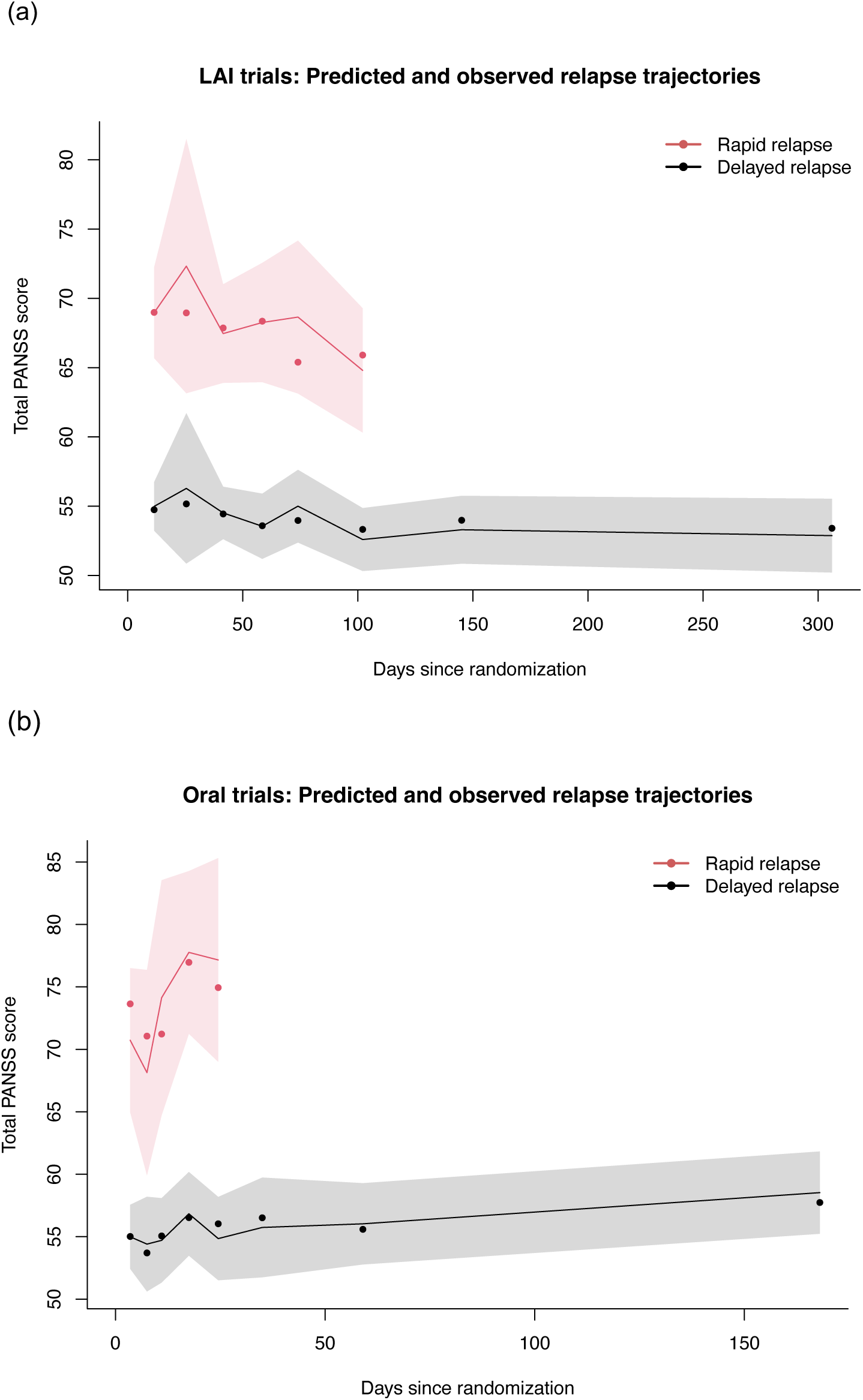
Predicted relapse trajectories in the two latent trajectory classes. a) Long-acting injectable (LAI) paliperidone palmitate trials. b) Oral paliperidone palmitate trials. Dots represent the weighted subject-specific predicted values of each latent trajectory class. The confidence interval range of observed values are shown in the shaded region.

### Latent class mixed modelling

In LAI trials, 224 of 271 participants (82.7%) had a relapse trajectory characterized by generally stable symptoms, with lower severity (Table 1). This latent class had a median time to relapse of 99 days, hence defined as ‘delayed relapse’. 47 participants (17.3%) had a relapse trajectory characterized by relatively severe psychotic symptoms from trial onset, with greater fluctuation of symptom severity. This latent class had a median time to relapse of 56 days, defined as ‘rapid relapse’.

**Table 1.** Baseline characteristics by relapse trajectory classes.

| **Long-Acting Injectable (LAI) Formulation** |  |  |  |  |
| --- | --- | --- | --- | --- |
| **Characteristics** | **Total cohort**<br>**(n = 271)** | **Delayed relapse**<br>**(n = 224)** | **Rapid relapse**<br>**(n = 47)** | **p-value** |
| Treatment group: |  |  |  | 0.119 |
| Discontinuation | 197 (72.7%) | 158 | 39 |  |
| Active treatment | 74 (27.3%) | 66 | 8 |  |
| Time to relapse<sup>a</sup> | 84 (88) | 99 (100) | 56 (50) | < 0.001 |
| Trial study: |  |  |  | 0.206 |
| Berwaerts (3-monthly) | 56 (20.7%) | 42 | 14 |  |
| Fu (1-monthly) | 82 (30.3%) | 68 | 14 |  |
| Hough (1-monthly) | 133 (49.1%) | 114 | 19 |  |
| Male sex | 155 (57.2%) | 127 (56.7%) | 28 (59.6%) | 0.841 |
| Age at screening<sup>b</sup> | 38.4 (11.2) | 38.2 (11.2) | 39.2 (11.4) | 0.596 |
| Baseline PANSS total<sup>b</sup> | 54.8 (13.7) | 52.7 (11.0) | 64.9 (19.6) | < 0.001 |
| Baseline CGI<sup>b</sup> | 2.70 (0.77) | 2.63 (0.76) | 3.02 (0.71) | 0.001 |
| **Oral Formulation** |  |  |  |  |
| **Characteristics** | **Total cohort**<br>**(n = 146)** | **Delayed relapse**<br>**(n = 107)** | **Rapid relapse**<br>**(n = 39)** | **p-value** |
| Treatment group: |  |  |  | 0.949 |
| Discontinuation | 108 (74.0%) | 79 | 29 |  |
| Active treatment | 38 (26.0%) | 28 | 10 |  |
| Time to relapse<sup>a</sup> | 25.5 (48.8) | 43 (55.5) | 10 (9) | < 0.001 |
| Trial study: |  |  |  | 0.195 |
| Kramer | 75 (51.4%) | 51 | 24 |  |
| Rui | 71 (48.6%) | 56 | 15 |  |
| Male sex | 72 (49.3%) | 50 (46.7%) | 22 (56.4%) | 0.396 |
| Age at screening<sup>b</sup> | 34.8 (11.5) | 34.4 (11.6) | 36.1 (11.2) | 0.434 |
| Baseline PANSS total<sup>b</sup> | 52.9 (10.2) | 50.8 (9.68) | 58.6 (9.46) | < 0.001 |
| Baseline CGI<sup>b</sup> | 2.83 (0.75) | 2.68 (0.71) | 3.23 (0.71) | < 0.001 |

<sup>a</sup> Median (IQR); <sup>b</sup> Mean (SD)

In oral formulation trials, 107 of 146 participants (73.3%) had a relapse trajectory characterized by generally stable symptoms, with lower severity and median time to relapse of 43 days (‘delayed relapse’) (Table 1). 39 participants (26.7%) had a trajectory characterized by relatively severe symptoms from trial onset, with greater fluctuation of symptom severity and median time to relapse of 10 days (‘rapid relapse’).

### Relapse trajectories in discontinuation compared to active treatment groups

In the LAI relapse cohort, the proportion of individuals with a rapid relapse trajectory did not differ significantly between the discontinuation (19.8%) and the active treatment group (10.8%; *X*^2^=2.44, *p*=0.119, Table 1). The trajectory class distribution did not differ significantly between treatment groups in the 1-monthly and 3-monthly formulation cohorts (eTable 6-7). In the oral cohort, rapid relapse was seen in 26.9% of discontinuation relapses, and 26.3% of the active treatment relapses (*X*^2^=0.004, *p*=0.949, Table 1).

### Symptom profiles at the time of relapse

For the LAI cohort, individuals with a rapid relapse trajectory had significantly higher scores on all PANSS items at time of relapse, except for 3 general psychopathology subscale items (Figure 2a). For the oral formulation cohort, the rapid relapse trajectory class had significantly more severe symptoms in all except 10 PANSS items at relapse (Figure 2c).

**Figure 2.**
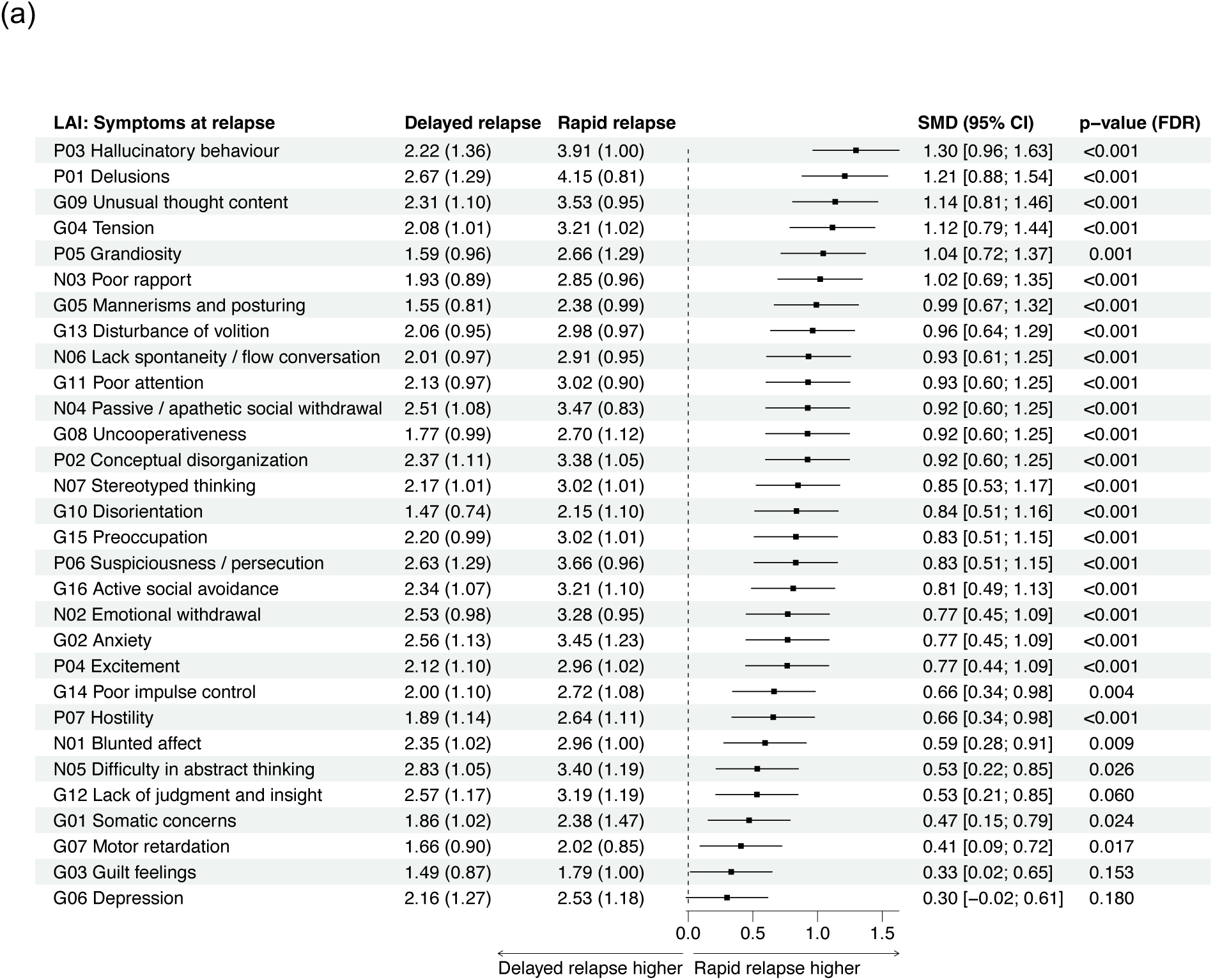
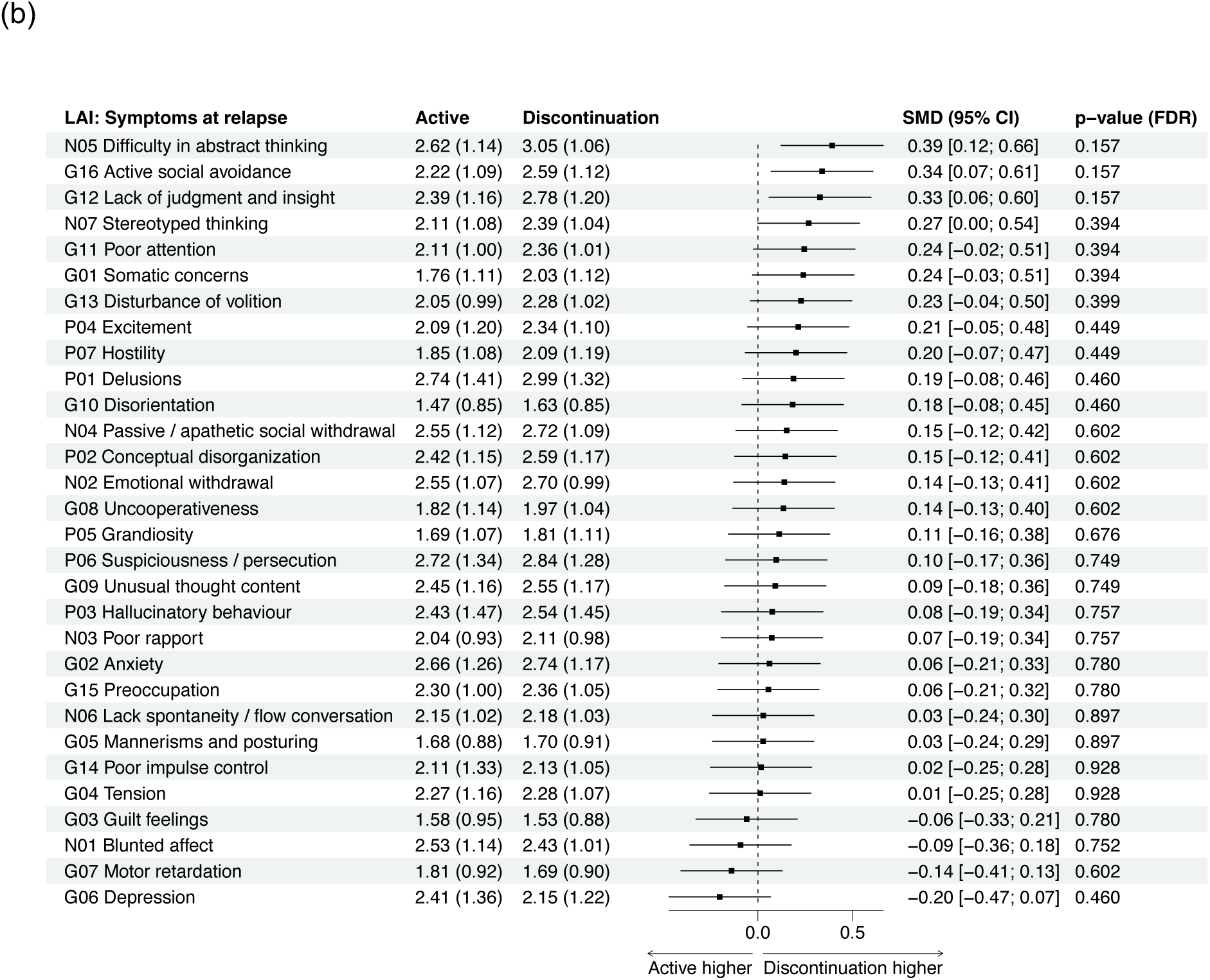
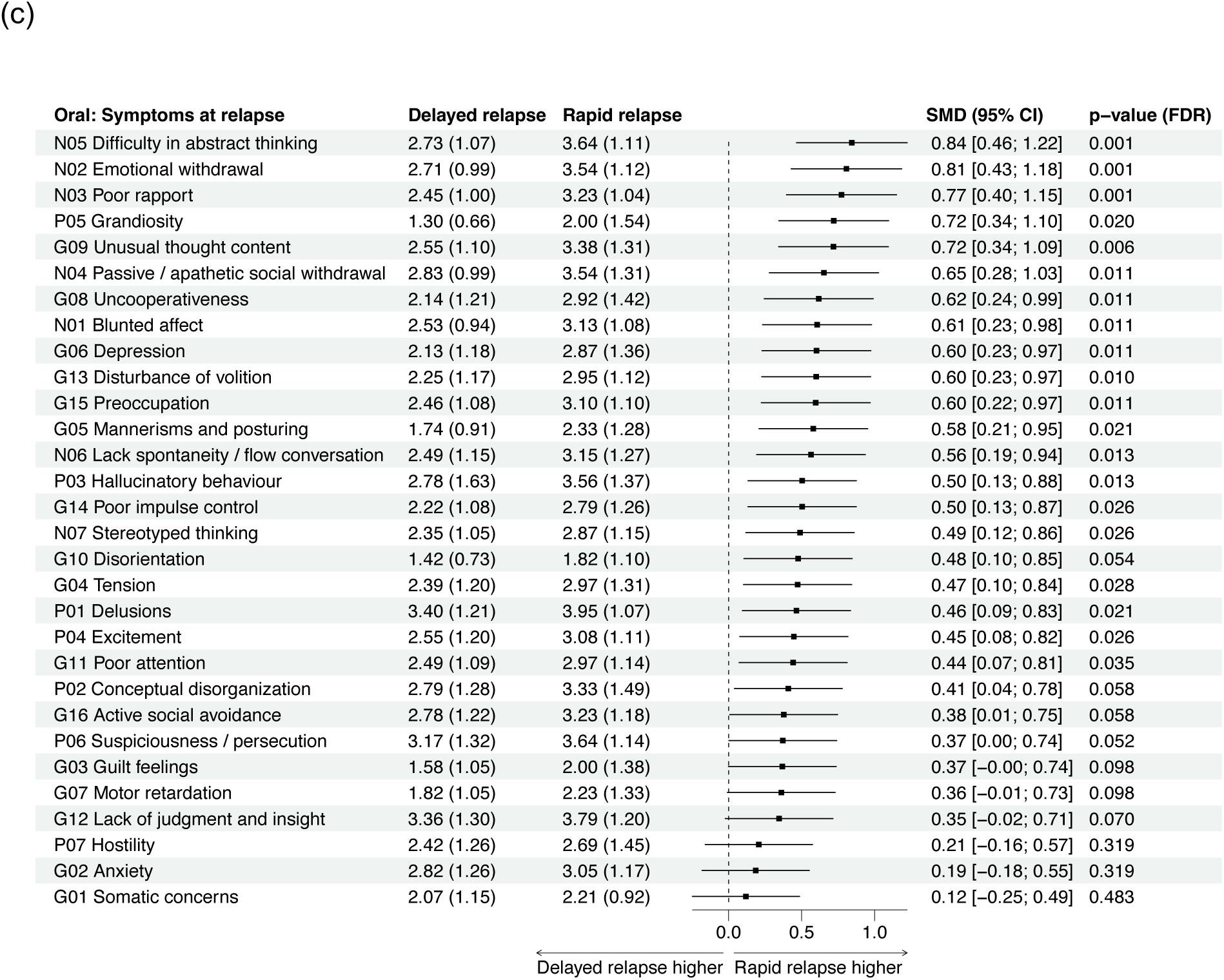
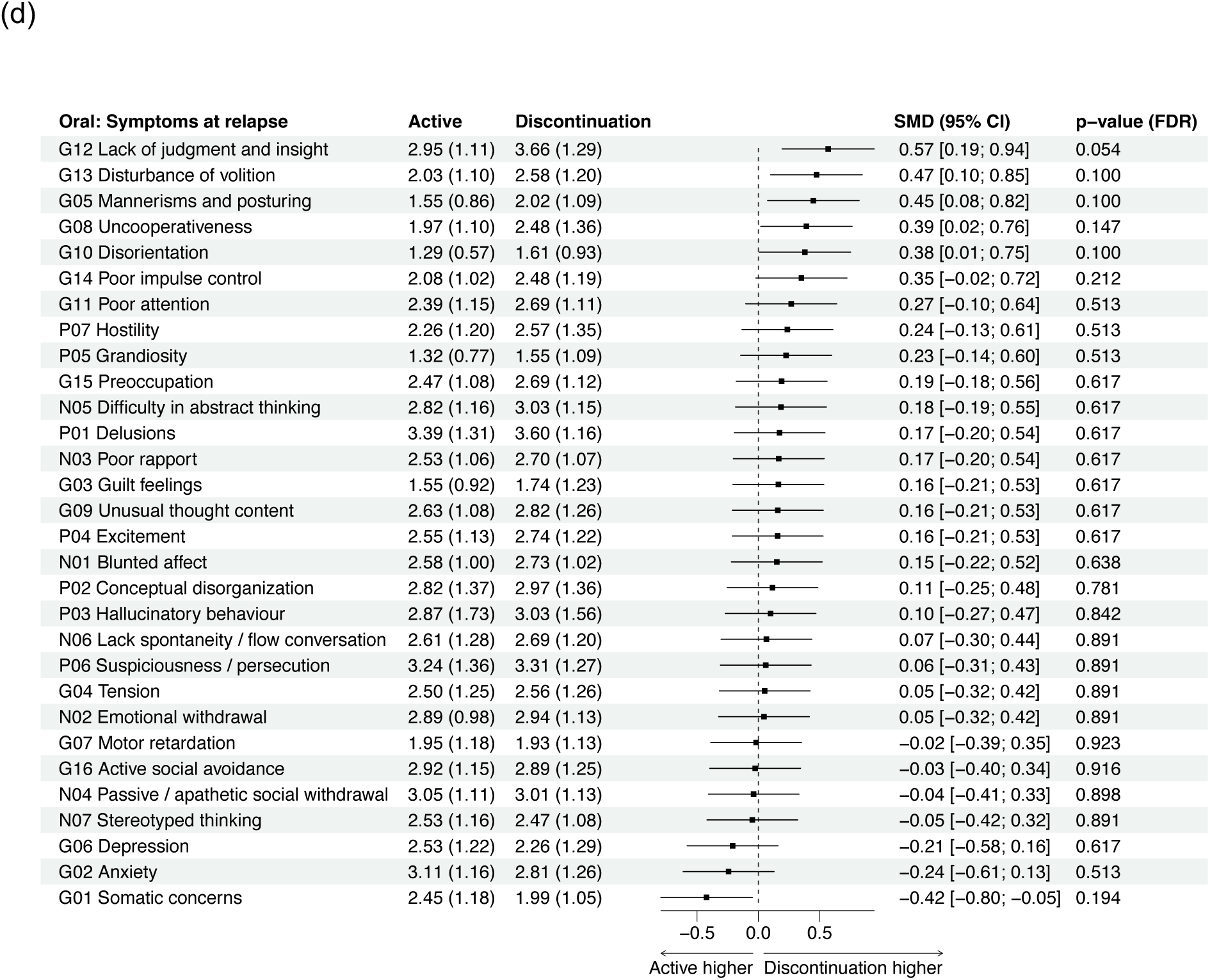
Forest plot of symptoms at the time of relapse. (a) Long-acting injectable (LAI) paliperidone palmitate trials by relapse trajectory class (b) Long-acting injectable (LAI) paliperidone palmitate trials by treatment group (c) Oral paliperidone palmitate trials by relapse trajectory class (d) Oral paliperidone palmitate trials by treatment group Mean (SD) scores are shown per class/group. Symptoms are ordered by effect size. *P*-values of *t*-tests are false discovery rate (FDR) corrected. SMD *standardized mean difference.* P *PANSS positive subscale.* N *negative subscale.* G *general psychopathology subscale*.

In both formulations, the discontinuation and active treatment groups did not differ significantly in symptom profiles at the time of relapse, and this includes symptoms previously suggested as indicative of withdrawal (i.e. agitation, anxiety, and depression) (Figure 2b, 2d).

### Baseline symptom profiles at the time of randomization

For the LAI cohort, the rapid relapse trajectory class had significantly higher baseline scores in terms of total PANSS (*p*<0.001), and 15 individual PANSS items compared to the delayed relapse class, across positive, negative and general symptom domains (Figure 3a). Baseline CGI severity was also significantly increased (*p*=0.001).

**Figure 3.**
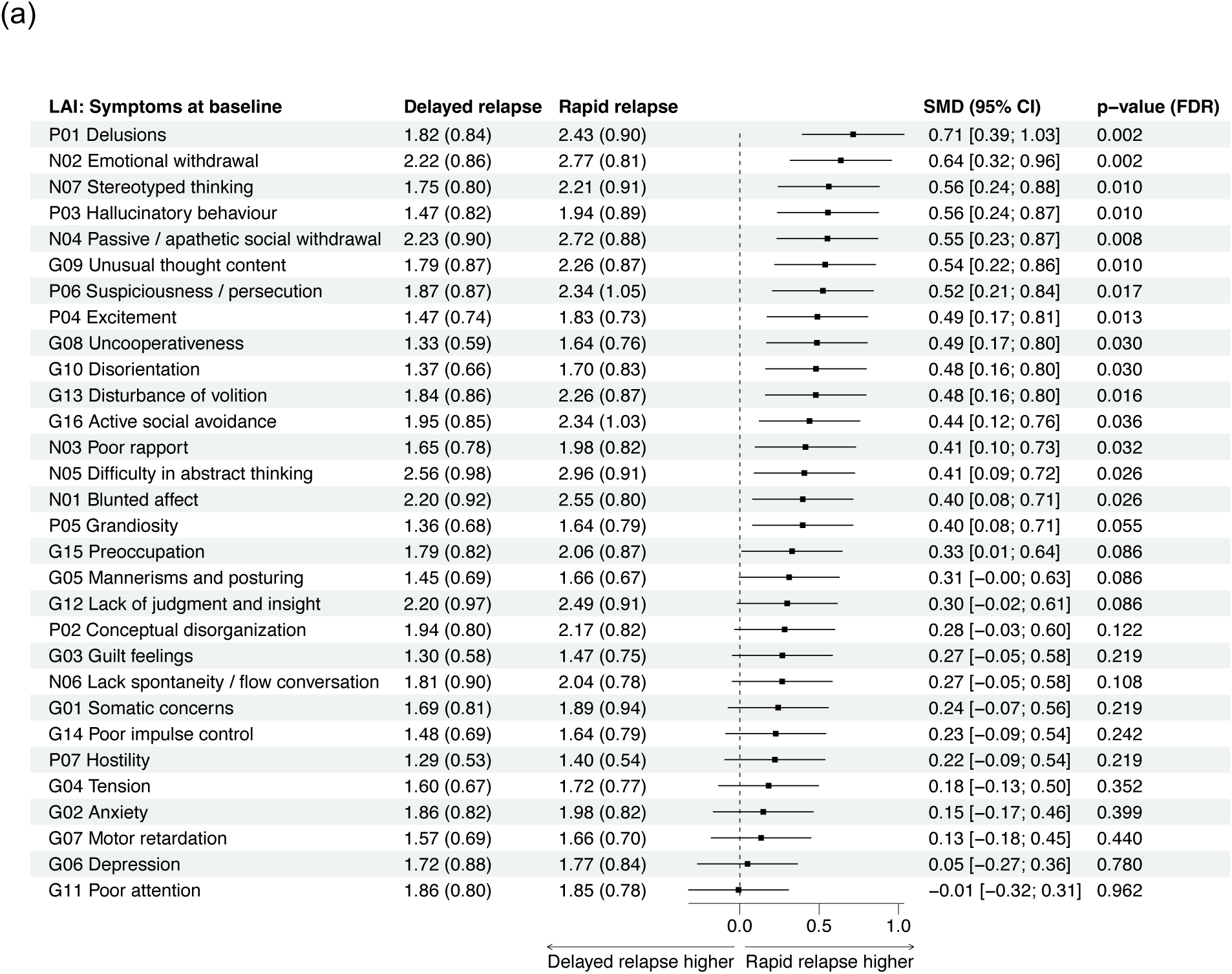
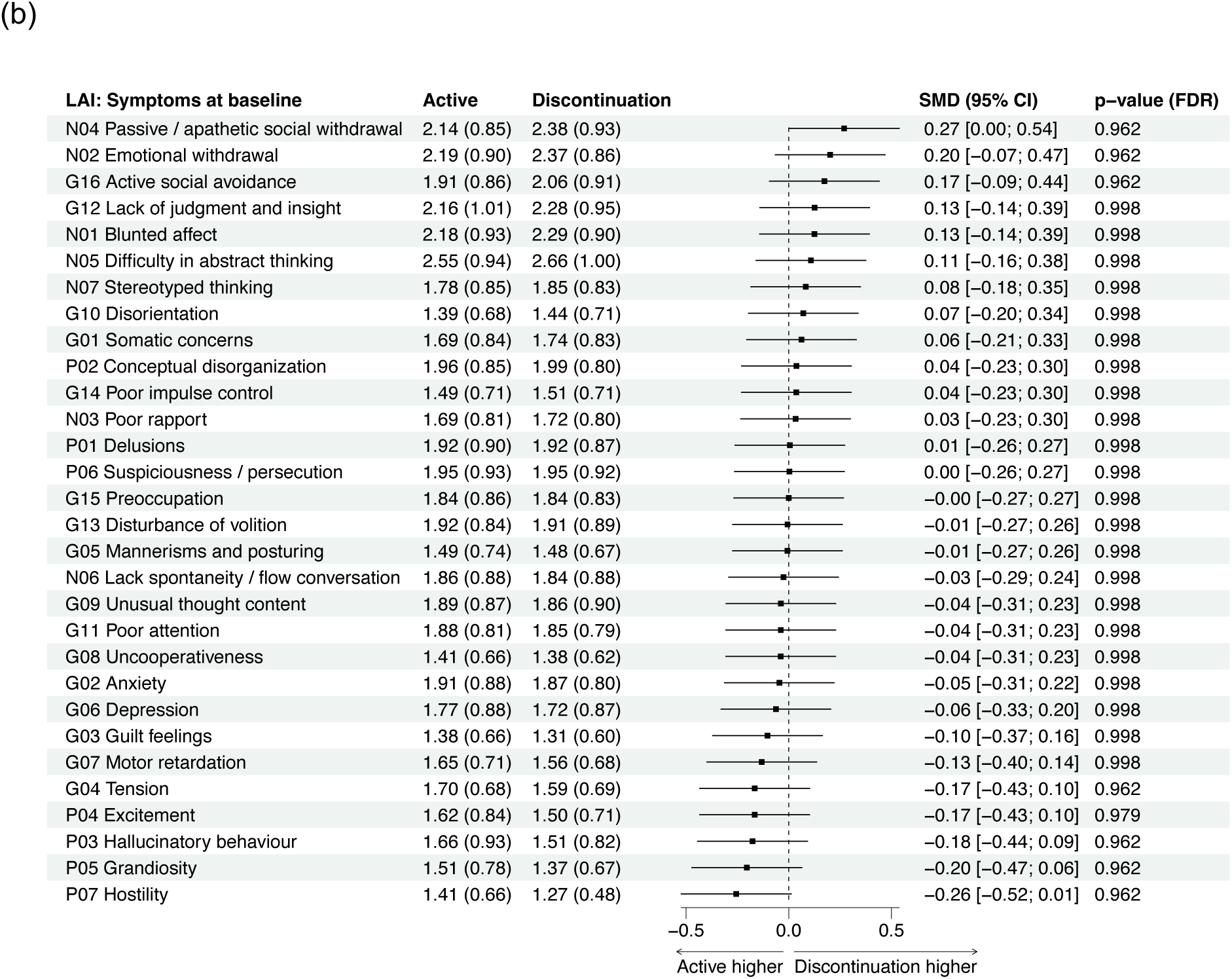
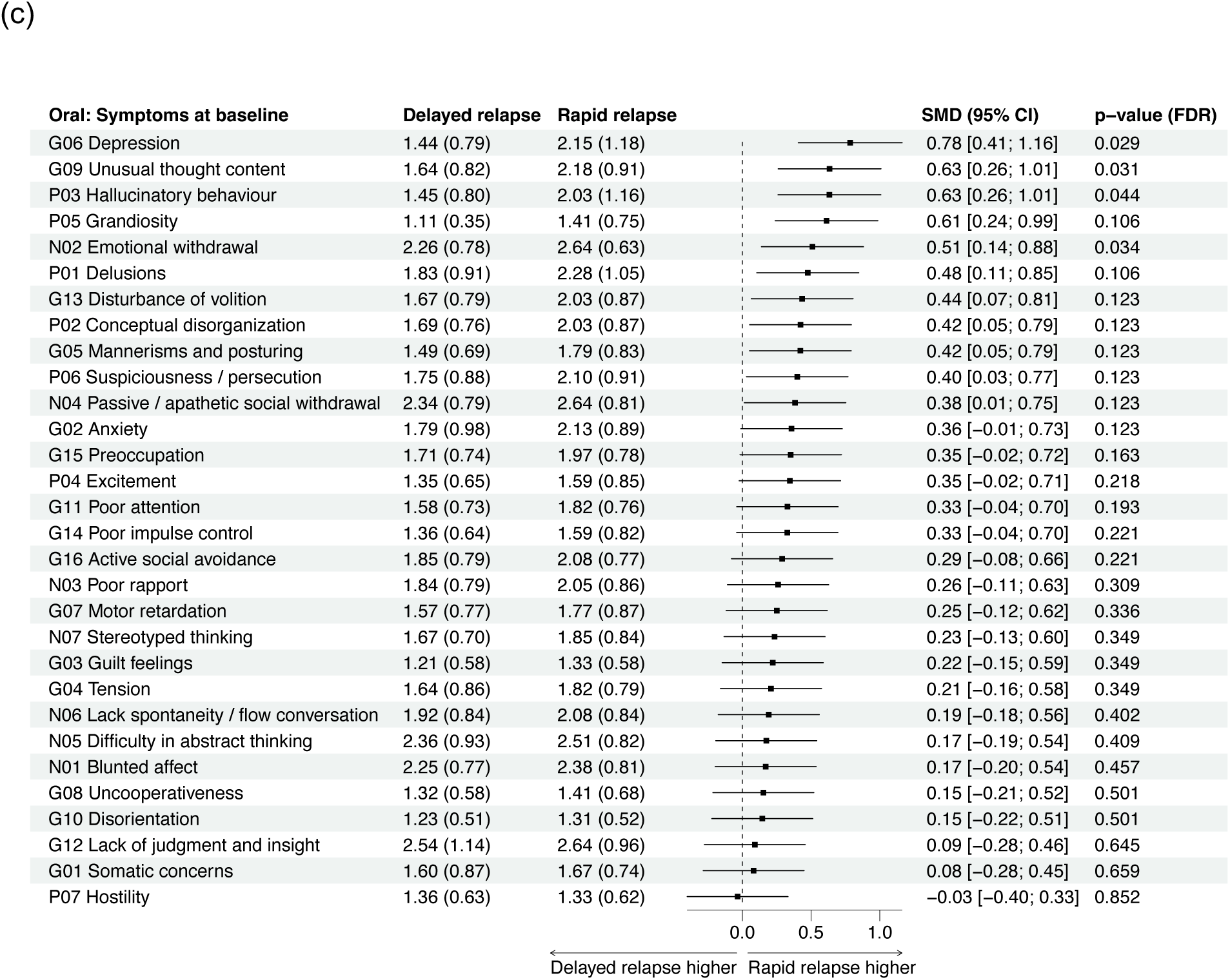
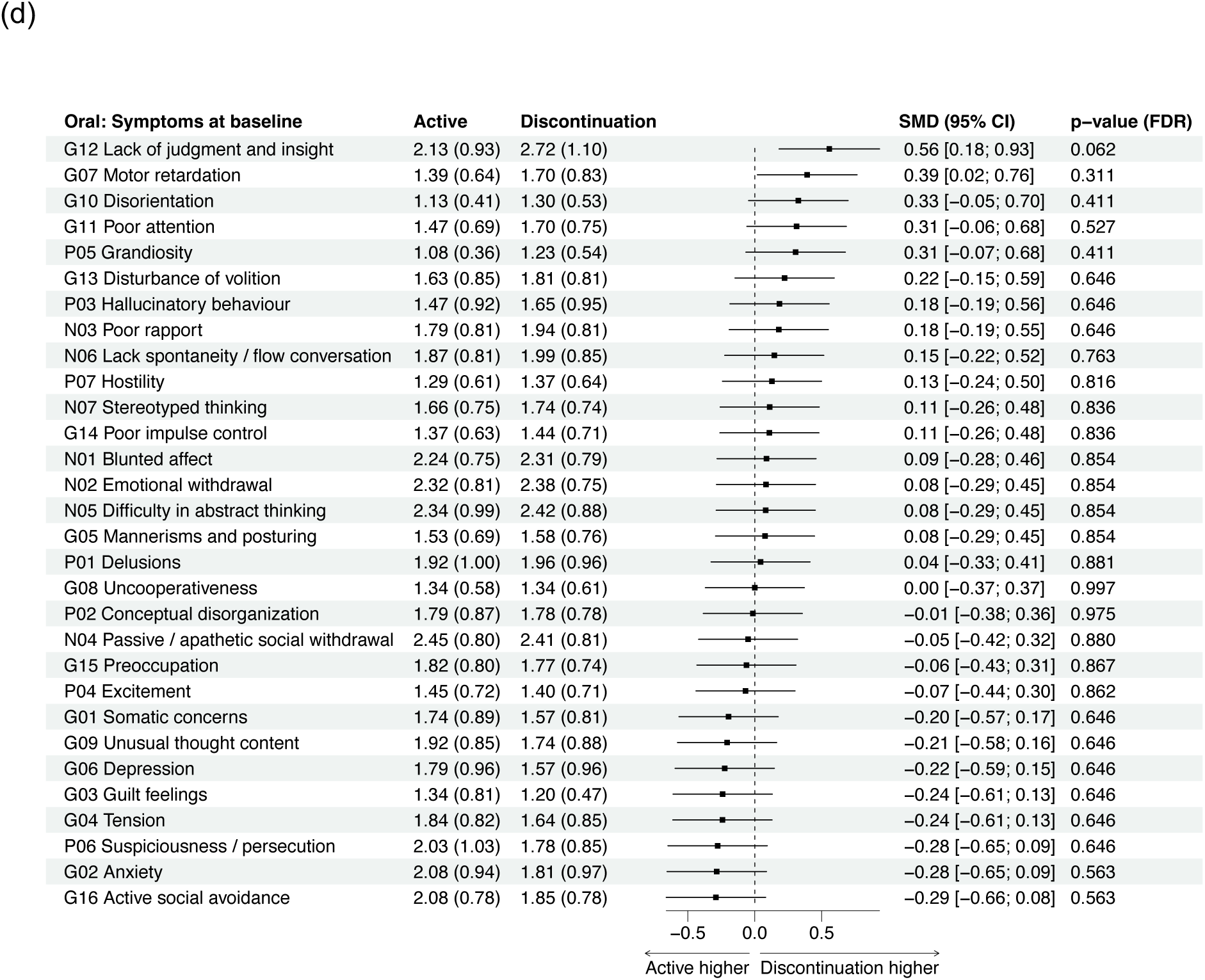
Forest plot of baseline symptoms at the time of randomization. (a) Long-acting injectable (LAI) paliperidone palmitate trials by relapse trajectory class (b) Long-acting injectable (LAI) paliperidone palmitate trials by treatment group (c) Oral paliperidone palmitate trials by relapse trajectory class (d) Oral paliperidone palmitate trials by treatment group Mean (SD) scores are shown per class/group. Symptoms are ordered by effect size. *p*-values of *t*-tests are false discovery rate (FDR) corrected. SMD *standardized mean difference.* P *PANSS positive subscale.* N *negative subscale.* G *general psychopathology subscale*.

In the oral formulation cohort, the rapid relapse trajectory class had significantly higher baseline total PANSS scores (*p*<0.001) and 4 individual PANSS items compared to the delayed relapse class (Figure 3c). Baseline CGI severity was also significantly increased (*p*<0.001).

In both formulations, the relapse trajectory classes did not differ significantly in study cohort, sex or age (Table 1).

## Discussion

Using LCMM we identified two distinct trajectory classes across 417 participants who experienced relapse in randomized double-blind discontinuation trials of paliperidone. A small group of participants experienced relapse with rapid onset, while the majority relapsed at a relatively delayed time point. The rapid relapse class was not over-represented in the treatment discontinuation group. This suggests that cases of rapid relapse observed in antipsychotic discontinuation trials are unlikely to be driven by withdrawal, and does not support the hypothesis that withdrawal-related mechanisms mediate a significant proportion of relapses observed in clinical trials. If acute withdrawal were the primary mechanism underlying rapid relapse, one would expect significantly greater proportions experiencing rapid relapse over delayed relapse in those who discontinued treatment. However, rapid relapse occurred independent of treatment status, supporting the hypothesis that pharmacological effects of acute withdrawal alone cannot explain the rapid relapses observed in antipsychotic discontinuation trials. This finding is contrary to the prediction that withdrawal-driven relapses should be more commonly observed after abrupt discontinuation. It is, however, consistent with meta-analyses showing no difference in relapse risk between abrupt and gradual antipsychotic discontinuation,^5,18^ and with recent clinical trials that have tested a gradual tapering approach.^34,35^

A plausible explanation, for the fact that the proportion of rapid relapses was similar in both continuation and discontinuation arms, is baseline symptom score deflation during trial enrolment. The pressure to meet eligibility thresholds, i.e. to be relatively symptom free at trial entry, may result in a marginal number of participants for study entry being downrated in symptom severity during baseline screening. There is evidence in the LAI and oral trials to support this interpretation, as those who eventually experienced rapid relapse had significantly higher severity scores at baseline (time of randomization), suggesting severity scores may have been slightly deflated to get just below a threshold, and allow inclusion into the trial. Following randomization, with rating constraints relieved, symptom assessments likely reflected true clinical status, giving the appearance of abrupt deterioration. Importantly, this pattern occurred in both discontinuation and active treatment groups, which is inconsistent with withdrawal-based hypotheses that predict rapid relapse specifically following discontinuation. This suggests that trial design and rating practices may influence observed relapse patterns, highlighting the importance of distinguishing genuine symptom recurrence from artefacts introduced during recruitment and assessment procedures in clinical trials.

Our analysis did not find evidence supporting a distinct withdrawal symptom profile in individuals that discontinued antipsychotic treatment. Withdrawal-driven relapse has been cited as potential confounds in classifying relapse in clinical trials, particularly after abrupt discontinuation.^36,37^ Yet, in the present analysis, symptom profiles at the time of relapse did not reveal a consistent pattern of elevated withdrawal-like symptoms among individuals that abruptly discontinued medication. There were no differences in symptoms at time of relapse between active treatment and discontinuation across both LAI and oral formulations, contradicting the hypothesis that relapse following antipsychotic discontinuation is commonly driven by pharmacologically mediated withdrawal syndromes. This finding also aligns with prior investigations of the same set of discontinuation RCTs, which demonstrated the risk of relapse is associated with overall levels of D_2_ receptor occupancy rather than rate of receptor occupancy decline,^19^ and with recent clinical trial evidence that tapering speed does not affect risk of relapse,^35^ strengthening the hypothesis that rapid relapses observed in discontinuation trials are a distinct phenomenon from an acute withdrawal reaction.

The finding that rapid relapsers had higher severity across a wide range of psychotic symptoms could suggest that individuals that experience rapid relapse had a more unstable disease course and greater susceptibility to abrupt clinical deterioration. This interpretation is supported by the fact that in both LAI and oral trials, rapid relapsers had a higher baseline CGI severity and higher baseline total PANSS scores than delayed relapsers.

Given the variability across studies in trial populations, inclusion and stabilization criteria, and definition of relapse, caution should be taken to avoid overinterpreting symptom profile differences. While relapse overall is markedly more common following antipsychotic discontinuation, our findings indicate rapid relapse can occur regardless of treatment status and appears to reflect an underlying vulnerability shaped by baseline severity and symptom stability.

These findings highlight the importance of interpreting rapid relapse cautiously in the clinic and not automatically attributing it to withdrawal syndrome or the patient having stopped treatment. Yet, this does not preclude the possibility that withdrawal-associated relapse occurs in specific clinical contexts, such as for patients with prolonged exposure to high-dose antipsychotics with abrupt cessation.^21,24^ or people abruptly stopping clozapine.

These findings also reinforce the need for careful patient monitoring of early warning signs and relapse risk assessment, rather than specific antipsychotic dose tapering schedules. Although gradual dose reduction may not directly lower risk of rapid relapse, it may facilitate earlier detection of clinical deterioration. Antipsychotic discontinuation should be informed by factors such as past treatment response and history of relapse. Considerations of the discontinuation method, baseline severity, duration of remittance and patient expectations should be embedded into guidelines and clinical care pathways for evidence-based de-prescribing.

Strengths of this study include the use of double-blind RCT data; this being the first analysis to systematically compare relapse trajectories between antipsychotic continuation and discontinuation groups; and the consistency of findings across both long-acting injectable and oral formulations. We note several limitations in our analysis. First, RCT participants are typically more stable and treatment adherent, with fewer comorbidities than real-world clinical populations. The present analysis was constrained by availability of key covariates (e.g. history of relapse and antipsychotic use) which may be important predictors of trajectory class membership.^38^ Future studies could apply predictive modelling approaches to identify factors associated with rapid relapse. Second, while LCMM is a powerful data-driven method that can identify latent trajectory classes without *a priori* assumptions, model selection can be subjective and there is a risk of overfitting, especially with smaller sample sizes. To mitigate this, we applied the best practice model selection procedure and validated class stability by iteratively fitting multiple models.^26,31^ Third, the original study trials were not designed to assess withdrawal phenomena. As there are currently no available validated measures of antipsychotic withdrawal symptoms, inferences about withdrawal-related symptoms are indirect. We mitigated this by analyzing PANSS items for known withdrawal-like features (e.g., anxiety, agitation, depression). Finally, as the included trials were of risperidone/paliperidone, it would be useful to determine if our relapse trajectories are seen following withdrawal from other antipsychotic agents. Future research incorporating multimodal data on receptor occupancy and plasma level measurements could help further clarify the role of pharmacological withdrawal in psychosis relapse.

In conclusion, this study identified two distinct trajectories to relapse (a rapid and delayed relapse trajectory) in randomized discontinuation trials of oral and LAI antipsychotics. Crucially, the rapid relapse trajectory class was not overrepresented in those that discontinued treatment, suggesting relapse mediated by pharmacological withdrawal is not a common feature after discontinuation. There was no symptom profile uniquely associated with withdrawal syndrome in those who discontinued medication. Symptom profiles at relapse were more severe in rapid relapsers, with a wide range of psychotic symptoms. Higher baseline severity predicted trajectory class (rapid relapse) in both cohorts, implicating biological mechanisms. These findings challenge the assumption that rapid relapse observed in clinical trials reflects a distinct withdrawal syndrome. Instead, rapid relapse is more likely to be linked to both trial factors and pre-existing biological vulnerability. Clinically, our data supports discontinuation strategies grounded in careful monitoring and risk assessment, rather than standardized pharmacological tapering schedules.

## Data Availability

All data produced in the present study are available upon reasonable request to the authors

## Acknowledgements

This study, carried out under YODA Project #2025-0740, used data obtained from the Yale University Open Data Access Project, which has an agreement with JANSSEN RESEARCH & DEVELOPMENT, L.L.C.. The interpretation and reporting of research using this data are solely the responsibility of the authors and does not necessarily represent the official views of the Yale University Open Data Access Project or JANSSEN RESEARCH & DEVELOPMENT, L.L.C.. The original proposal can be found: https://yoda.yale.edu/data-request/2025-0740/.

RAMs work is funded by a Wellcome Trust Clinical Research Career Development Fellowship (224625/Z/21/Z) and is supported by the NIHR Oxford Health Biomedical Research Centre. TP is supported by a Wellcome Trust Early Career Fellowship, Maudsley Charity, the Brain and Behaviour Research Foundation, the UK Academy of Medical Sciences, and the UK Research and Innovation Hub for Metabolic Psychiatry (grant reference MR/Z503563/1, platform grant code MR/Z000548/1). The views expressed are those of the author(s) and not necessarily those of the NIHR or the Department of Health and Social Care.

