## Supplementary Materials for "The Course of Relapse Following Antipsychotic Discontinuation in Schizophrenia: A Test of the Antipsychotic Withdrawal Syndrome Hypothesis of Relapse"

**Supplementary Material**

*Supplementary Material 1: Details of LCMM model fit and statistical analysis*

The LCMM model fit was conducted iteratively in the following manner. First, the simplest linear transformation model of total PANSS score varying with days since randomization up to relapse was fit. Then, a model with the rescaled cumulative distribution function (CDF) of a Beta distribution was fit to assess potential non-linear trajectories.[1] Next, random effects were added to account for heterogeneity in symptom trajectories within latent classes. Finally, models with 1, 2, 3 and 4 latent classes were estimated and compared. To maximize clinical meaningfulness of the latent trajectory classes, models in which the smallest class had less than 5% of the total participants were excluded.

The optimal model with the most appropriate number of latent classes was determined according to the following criteria:

1. Size-adjusted Bayesian Information Criterion (BIC): lower values indicate better model fit[2]
2. Entropy: describes class discriminability, with values closer to 1 indicating less classification uncertainty and better distinction between classes[3] – values greater than 0.5 is regarded acceptable[4]
3. Posterior probability of classification: average latent class probabilities for most likely latent class membership – higher probabilities indicate better classification, with 70%+ regarded as acceptable[4]

The non-linear Beta distribution models did not converge, and the models without random effects had higher BIC and lower entropy than the models with random effects.

For all between group symptom comparisons, the appropriate statistical test was applied: Shapiro-Wilk tests were conducted before applying Welch's *t*-test for continuous normal variables and Mann-Whitney *U* test for continuous non-normal variables. The Cohen's *d* standardized mean difference (SMD) and the standard error (SE) of the SMD was computed. Given that 30 tests were made for each group comparison, the false discovery rate (FDR) approach was implemented to account for multiple comparison testing.[5] For completeness, both the 95% confidence interval of SMD and the FDR-corrected *p*-value were computed. The SMD was selected as the effect size measure for better comparability between symptoms and study trials. Subgroup comparisons between 1-monthly and 3-monthly LAI trials were not conducted due to the small sample sizes.

Threshold of statistical significance was set at 0.05 (two-tailed, and FDR corrected where appropriate). Error bars represent the 95% confidence interval unless otherwise stated. All statistical analyses were conducted in R version 4.3.0[6] on the secure YODA platform[7] using R packages lcmm v.2.1.0[1], compareGroups v.4.7.2[8] and forestploter v.1.1.3[9].

*Supplementary Material 2: Extended results*

In the LCMM model fit analyses, the 2-class model had the lowest size-adjusted BIC and highest entropy. The 4-class models had latent classes with less than 5% participants. The average latent class probabilities for most likely class membership were acceptable for all classes in the 2-class model (LAI: delayed 89.5%, rapid 76.7%; oral: delayed 96.7%, rapid 86.3%).

Model fit for the LAI trials split by 1-monthly and 3-monthly formulation showed a similar trend to the combined LAI cohort analysis. The 2-class linear models with random effects showed the best model fit. The average latent class probabilities for most likely class membership were acceptable in all classes for the 2-class models (range: 76.6%-90.2%). The goodness-of-fit of the final 2-class models were acceptable.

In the secondary analysis of LAI formulations, 177 of 215 participants (82.3%) with 1-monthly LAI had the ‘delayed relapse’ trajectory, with a median time to relapse of 96 days. The remaining 38 participants (17.7%) had the ‘rapid relapse’ trajectory with median 56 days to relapse. 41 of 56 participants (73.2%) in the 3-monthly LAI trial had a ‘delayed relapse’ trajectory with median 116 days to relapse, while 15 participants (26.8%) had a ‘rapid relapse’ trajectory with median 63 days to relapse.

For baseline differences in the two LAI formulations, the baseline total PANSS score ($p$ = 0.006) in the 1-monthly LAI cohort was significantly higher in the rapid relapse trajectory class, but only marginally higher in the 3-monthly LAI cohort ($p$ = 0.050). The baseline total CGI score was significantly higher in the rapid relapse class for the 3-monthly LAI cohort ($p$ = 0.009), but not for the 1-monthly LAI cohort ($p$ = 0.235). In both LAI formulations, there were no significant differences between treatment groups in any of the covariates.

As expected, given the randomized nature of the studies, there were no significant differences in baseline symptom profiles between discontinuation and active treatment groups (Figure 3b, 3d, eTable4).

*Supplementary Material 3: Relapse criteria in the original studies*

**<u>Fu:</u> any of the following —**

1. Psychiatric hospitalization
2. Any intervention employed to avert imminent hospitalization due to worsening of symptoms (e.g. increase in the level of psychiatric care) or the need for additional antipsychotics, antidepressant or mood stabilizers
3. Clinically significant self-injury, suicidal or homicidal ideation, violent behaviours
4. Total PANSS score increase for 2 consecutive visits by:
    a. If baseline score > 45: increase by 25% from baseline
    b. If baseline score ≤ 45: 10-point increase
5. Increase in CGI-Severity score for 2 consecutive visits to:
    a. If baseline score ≤ 3: increase in 2 or more points
    b. If baseline score ≥ 4: increase in 1 or more points
6. Worsening in specific PANSS items (to ≥6 if baseline ≤ 4 or to ≥5 if baseline ≤3):
    a. P01 Delusions
    b. P02 Conceptual disorganization
    c. P03 Hallucinatory behaviour
    d. P04 Excitement
    e. P06 Suspiciousness / persecution
    f. P07 Hostility
    g. G08 Uncooperativeness
    h. G14 Poor impulse control

**<u>Hough, Berwaerts, Kramer, Rui:</u> any of the following —**

1. Psychiatric hospitalization (involuntary or voluntary admission)
2. Increase in the level of psychiatric care (e.g. from clinic visits to day treatment)
3. Total PANSS score increase for 2 consecutive visits by:
    a. If baseline score > 40: increase by 25% from baseline
    b. If baseline score ≤ 40: 10-point increase
4. Increase in CGI-Severity score for 2 consecutive visits to:
    a. If baseline score ≤ 3: increase to 4
    b. If baseline score = 4: increase to 5
5. Deliberate self-injury
6. Suicidal or homicidal ideation
7. Clinically significant aggressive behaviour
8. Increase in specific PANSS item scores for 2 consecutive assessments:
    a. P01 Delusions
    b. P02 Conceptual disorganization
    c. P03 Hallucinatory behaviour
    d. P06 Suspiciousness / persecution
    e. P07 Hostility
    f. G08 Uncooperativeness

*Supplementary Material 4: Baseline inclusion criteria in the original studies*

**Fu:** Patients with a diagnosis of either a schizophrenia or schizoaffective disorder for at least one year and with prominent mood symptoms at screening. Specifically, patients who were experiencing exacerbation of psychotic symptoms, defined as a score of ≥ 4 on 3 or more of specific PANSS items:

a. P01 Delusions
b. P02 Conceptual disorganization
c. P03 Hallucinatory behaviour
d. P04 Excitement
e. P06 Suspiciousness/persecution
f. P07 Hostility
g. G04 Tension
h. G08 Uncooperativeness
i. G14 Poor impulse control

**Hough and Berwaerts:** Patients with a diagnosis of schizophrenia for at least one year. Total PANSS score under 120 (no minimum score required). Not treatment resistant.

**Kramer and Rui:** Patients with a diagnosis of schizophrenia for at least one year. Total PANSS score between 70 and 120. Not treatment resistant.

### Supplementary Tables

#### eTable 1. Proposed antipsychotic withdrawal symptoms compared to relapse

| Class | Onset | Duration | Symptoms |
|---|---|---|---|
| Withdrawal | Within days after drug discontinuation (peak onset of 36 to 96 hours) | 6 weeks or more depending on drug half-life | New symptoms common to CNS drugs: nausea, headache, insomnia, tremor, anxiety, irritability, agitation, aggression, inattention, dysphoria, depression<br><br>Serotonergic withdrawal syndrome: flu-like symptoms, dizziness, diarrhoea, confusion, disorientation, restlessness/clonus<br><br>Adrenergic withdrawal syndrome: headache, anxiety, palpitation, increased blood pressure or heart rate, sweating, presyncope<br><br>Histaminic withdrawal syndrome: irritability, depressed affect, loss of appetite, increased inducible seizure, lethargy or amnesia<br><br>Muscarinic withdrawal syndrome: agitation, nausea, vomiting, abdominal cramp, hypothermia, tremor, parkinsonism, restlessness, insomnia |
| Relapse or recurrence | Any time post discontinuation | Days, weeks | Return of ongoing episodes or a new psychotic episode with symptoms known to the patient |

Adapted from Chouinard et al. (2017)[10]

#### eTable 2. Study design and participant characteristics

| Study | Follow-up | Relapse cases by intervention | Participant characteristics |
|---|---|---|---|
| Fu[11] | Up to 16 months | Discontinuation (*n*=57, 69.5%)<br>1-monthly LAI (*n*=25, 30.5%) | Schizophrenia or schizoaffective disorder experiencing exacerbation (48% male, mean age 36.3) |
| Hough[12] | Up to 16 months | Discontinuation (*n*=97, 72.9%)<br>1-monthly LAI (*n*=36, 27.1%) | Schizophrenia disorder (54% male, mean age 40.7) |
| Berwaerts[13] | Up to 16 months | Discontinuation (*n*=43, 76.8%)<br>3-monthly LAI (*n*=13, 23.2%) | Schizophrenia disorder (79% male, mean age 35.8) |
| Kramer[14] | Up to 12 months | Discontinuation (*n*=52, 69.3%)<br>Oral (*n*=23, 30.7%) | Schizophrenia disorder (56% male, mean age 37.9) |
| Rui[15] | Up to 14 months | Discontinuation (*n*=56, 78.9%)<br>Oral (*n*=15, 21.1%) | Schizophrenia disorder (42% male, mean age 31.8) |

**eTable 3. Model fit statistics for long-acting injectable (LAI) and oral trials**

| Model | BIC | Entropy | Proportion of individuals in each class | | | |
|---|---|---|---|---|---|---|
| **LAI trials (1-monthly and 3-monthly combined, *n* = 271)** | | | | | | |
| 1-class | 10396.27 | - | 100% | | | |
| 2-class | 10377.55 | 0.60 | 82.7% | 17.3% | | |
| 3-class | 10384.02 | 0.50 | 63.1% | 22.5% | 14.4% | |
| 4-class | 10395.60 | 0.60 | 40.2% | 39.1% | 15.5% | 5.2% |
| **Oral trials (*n* = 146)** | | | | | | |
| 1-class | 6581.00 | - | 100% | | | |
| 2-class | 6516.34 | 0.80 | 73.3% | 26.7% | | |
| 3-class | 6528.63 | 0.56 | 53.4% | 27.4% | 19.2% | |
| 4-class | 6543.58 | 0.47 | 50.7% | 27.4% | 21.9% | 0% |

**eTable 4. Baseline characteristics of long-acting injectable cohort by treatment group**

| ***Long Acting Injectable Formulation*** | | | | |
|---|---|---|---|---|
| **Characteristics** | Total cohort (*n* = 271) | Discontinuation (*n* = 197) | Active treatment (*n* = 74) | *p*-value |
| **Trajectory class:** | | | | 0.119 |
| **Delayed relapse** | 224 (82.7%) | 158 (80.2%) | 66 (89.2%) | |
| **Rapid relapse** | 47 (17.3%) | 39 (19.8%) | 8 (10.8%) | |
| **Time to relapse[a]** | 84 (88) | 85 (90) | 81.5 (78.5) | 0.451 |
| **Trial study:** | | | | 0.639 |
| **Berwaerts (3-monthly)** | 56 (20.7%) | 43 (21.8%) | 13 (17.6%) | |
| **Fu (1-monthly)** | 82 (30.3%) | 57 (28.9%) | 25 (33.8%) | |
| **Hough (1-monthly)** | 133 (49.1%) | 97 (49.2%) | 36 (48.6%) | |
| **Male sex** | 155 (57.2%) | 109 (55.3%) | 46 (62.2%) | 0.382 |
| **Age at screening[b]** | 38.4 (11.2) | 38.9 (11.4) | 36.9 (10.8) | 0.188 |
| **Baseline PANSS total[b]** | 54.8 (13.7) | 54.7 (13.0) | 55.0 (15.4) | 0.897 |
| **Baseline CGI[b]** | 2.70 (0.77) | 2.71 (0.74) | 2.66 (0.83) | 0.661 |
| ***Oral Formulation*** | | | | |
| **Characteristics** | Total cohort (*n* = 146) | Discontinuation (*n* = 108) | Active treatment (*n* = 38) | *p*-value |
| **Trajectory class:** | | | | 1.000 |
| **Delayed relapse** | 107 (73.3%) | 79 (73.1%) | 28 (73.7%) | |
| **Rapid relapse** | 39 (26.7%) | 29 (26.9%) | 10 (26.3%) | |
| **Time to relapse[a]** | 25.5 (48.8) | 25.5 (48.8) | 25.5 (45.5) | 0.976 |
| **Trial study:** | | | | 0.261 |
| **Kramer** | 75 (51.4%) | 52 (48.1%) | 23 (60.5%) | |
| **Rui** | 71 (48.6%) | 56 (51.9%) | 15 (39.5%) | |
| **Male sex** | 72 (49.3%) | 54 (50.0%) | 18 (47.4%) | 0.928 |
| **Age at screening[b]** | 34.8 (11.5) | 34.6 (11.5) | 35.4 (11.5) | 0.716 |
| **Baseline PANSS total[b]** | 52.9 (10.2) | 53.1 (10.1) | 52.3 (10.6) | 0.678 |
| **Baseline CGI[b]** | 2.83 (0.75) | 2.84 (0.71) | 2.79 (0.84) | 0.730 |

[a] Median (IQR); [b] Mean (SD)

#### eTable 5. Model fit statistics for LAI cohort split by formulation type

| Model | BIC | Entropy | Proportion of individuals in each class | | | |
|---|---|---|---|---|---|---|
| **3-monthly LAI formulation (*n* = 105)** | | | | | | |
| **1-class** | 2554.65 | - | 100% | | | |
| **2-class** | 2561.52 | 0.59 | 73.2% | 26.8% | | |
| **3-class** | 2573.59 | 0.29 | 66.1% | 33.9% | 0% | |
| **4-class** | 2585.67 | 0.23 | 60.7% | 39.3% | 0% | 0% |
| **1-monthly LAI formulation (*n* = 215)** | | | | | | |
| **1-class** | 12374.21 | - | 100% | | | |
| **2-class** | 7818.75 | 0.57 | 82.3% | 17.7% | | |
| **3-class** | 7834.87 | 0.30 | 77.7% | 22.3% | 0% | |
| **4-class** | 7837.95 | 0.46 | 76.7% | 11.6% | 11.6% | 0% |

#### eTable 6. Characteristics of 3-monthly LAI cohort by trajectory class

| Characteristics | Total cohort (*n* = 56) | Delayed relapse (*n* = 41) | Rapid relapse (*n* = 15) | *p*-value |
|---|---|---|---|---|
| **Treatment group:** | | | | 0.730 |
| **Discontinuation** | 43 (76.8%) | 32 (78.0%) | 11 (73.3%) | |
| **Active treatment** | 13 (23.2%) | 9 (22.0%) | 4 (26.7%) | |
| **Time to relapse**[a] | 90.5 (78.8) | 116 (72) | 63 (42) | < 0.001 |
| **Male sex** | 44 (78.6%) | 30 (73.2%) | 14 (93.3%) | 0.149 |
| **Age at screening**[b] | 36.2 (11.3) | 37.1 (10.9) | 32.1 (12.1) | 0.072 |
| **Baseline PANSS total**[b] | 59.0 (16.1) | 55.5 (11.3) | 68.5 (22.9) | 0.050 |
| **Baseline CGI**[b] | 2.82 (0.61) | 2.73 (0.67) | 3.07 (0.26) | 0.009 |

[a] Median (IQR); [b] Mean (SD)

#### eTable 7. Characteristics of 1-monthly LAI cohort by trajectory class

| Characteristics | Total cohort (*n* = 215) | Delayed relapse (*n* = 177) | Rapid relapse (*n* = 38) | *p*-value |
|---|---|---|---|---|
| **Treatment group:** | | | | 0.366 |
| **Discontinuation** | 154 (71.6%) | 125 (70.1%) | 30 (78.9%) | |
| **Active treatment** | 61 (28.4%) | 53 (29.9%) | 8 (21.1%) | |
| **Time to relapse**[a] | 84 (88.5) | 96 (108) | 56 (51.5) | < 0.001 |
| **Male sex** | 111 (51.6%) | 94 (53.1%) | 17 (44.7%) | 0.448 |
| **Age at screening**[b] | 39.0 (11.2) | 38.5 (11.4) | 41.7 (9.82) | 0.074 |
| **Baseline PANSS total**[b] | 53.7 (12.8) | 52.2 (10.8) | 60.9 (17.9) | 0.006 |
| **Baseline CGI**[b] | 2.67 (0.80) | 2.63 (0.79) | 2.82 (0.87) | 0.235 |

[a] Median (IQR); [b] Mean (SD)

### Supplementary Figures

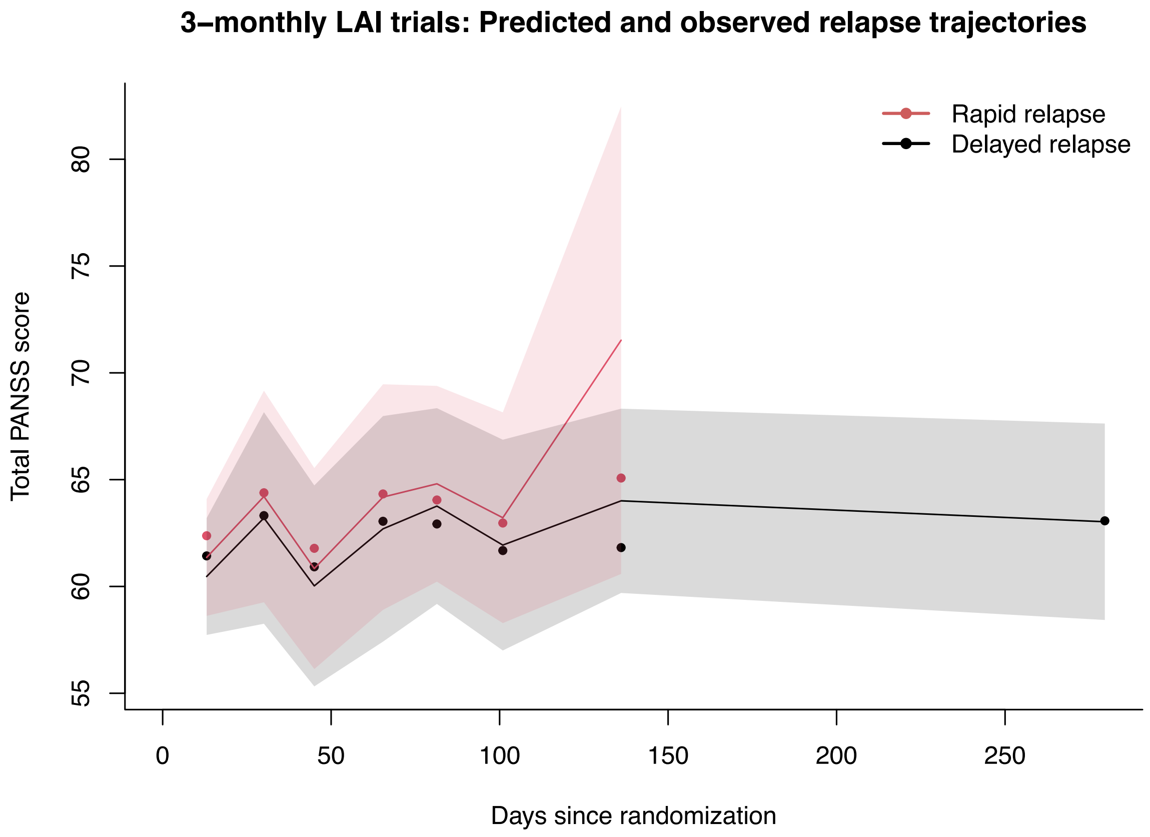


**eFigure 1. Predicted relapse trajectories with 2 latent trajectory classes in 3-monthly long-acting injectable paliperidone palmitate (LAI) trials.** Dots represent the weighted subject-specific predicted values. The confidence interval range of observed values are shown in the shaded region.

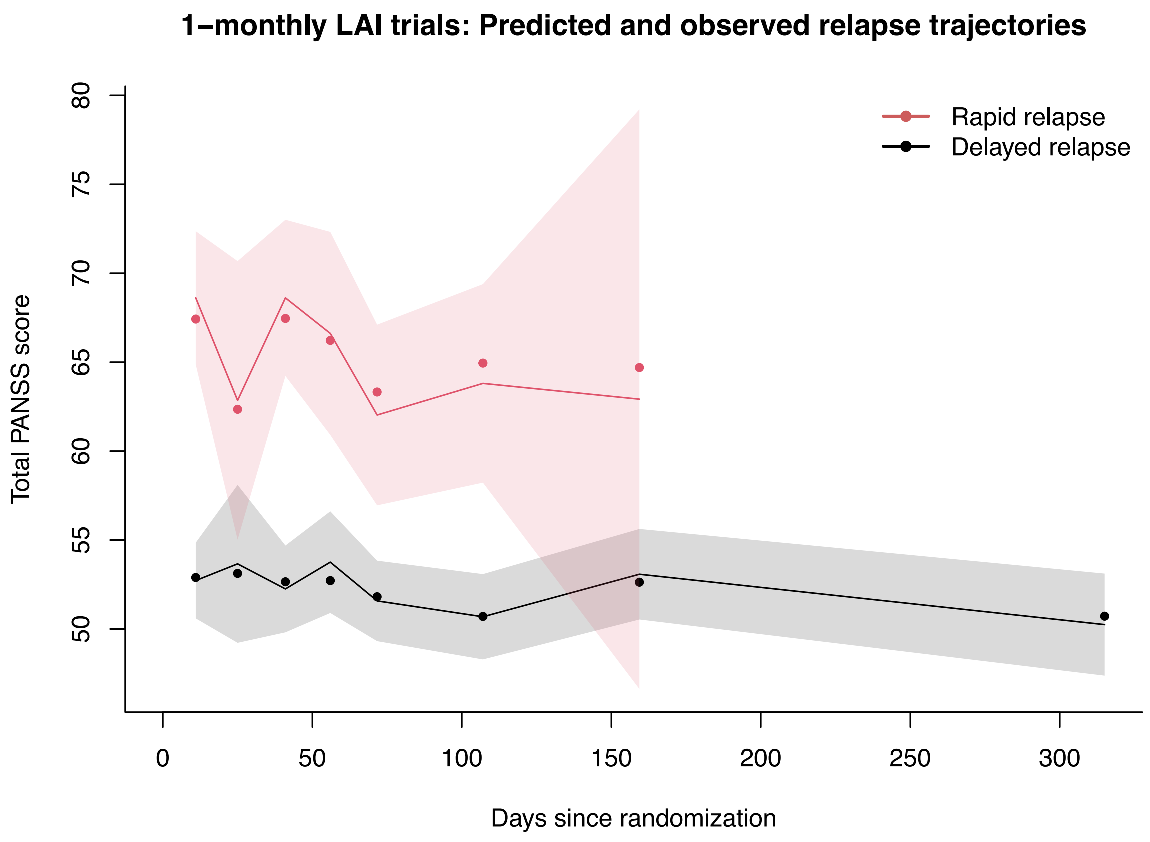


**eFigure 2. Predicted relapse trajectories with 2 latent trajectory classes in 1-monthly long-acting injectable paliperidone palmitate (LAI) trials.** Dots represent the weighted subject-specific predicted values. The confidence interval range of observed values are shown in the shaded region.
